# Analyses of academician cohorts generate biased pandemic excess death estimates

**DOI:** 10.1101/2024.03.20.24304645

**Authors:** John P.A. Ioannidis

## Abstract

**Objective:** Death data from cohorts of academicians have been used to estimate pandemic excess deaths. We aimed to evaluate the validity of this approach.

**Study design and setting:** Data were analyzed from living and deceased member lists from Mainland China, UK and Greece academies; and Nobel laureates (and US subset thereof). Samples of early elected academicians were probed for unrecorded deaths; datasets overtly missing deaths were excluded from further analyses. Actuarial risks were compared against the general population in the same country in respective age strata. Relative incidence risk increases in death in active pandemic periods were compared to population-wide pandemic excess death estimates for the same country.

**Results:** Royal Society and Academy of Athens datasets overtly missed deaths. Pre-pandemic death rates were 4-12-fold lower in the Chinese Academy of Engineering (CAE) versus respective age strata of the Mainland China population. A +158% relative increase in death risk was seen in CAE data during the first 12-months of wide viral spread. Both increases (+34% in British Academy) and decreases (-27% in US Nobel laureates) in death rates occurred in pandemic (2020-22) versus pre-pandemic (2017-2019) years; point estimates were far from known excess deaths in the respective countries (+6% and +14%, respectively). Published excess death estimates for urban-dwelling Mainland China selectively analyzed CAE, but not another Chinese academy (Chinese Academy of Sciences) with half the pandemic death rates.

**Conclusion:** Missingness, lack of representativeness, large uncertainty, and selective analysis reporting make data from academy rosters unreliable for estimating general population excess deaths.

## INTRODUCTION

Estimating pandemic excess death burden is difficult, even in countries with excellent death registration systems, because estimates depend on many assumptions.^1,2^ In countries with deficient death registration, one option is to make inferences from cohorts where death data are hopefully complete. One approach is to use cohorts of members of prestigious academies, where birth and death dates are recorded for their members. This approach was widely popularized when in 2023 a highly-read New York Times article^3^ noticed that deaths of Chinese Academy of Engineering (CAE) and Chinese Academy of Sciences (CAS) members peaked during December 2022 and January 2023. Linked to explosive spread of COVID-19 in Mainland China after removal of restrictive measures, deaths of academicians were believed to reflect the magnitude of excess death toll across Mainland China. This concept adopted in a meticulous epidemiological study concluded from CAE death data that the urban-dwelling population of Mainland China had 917,000 excess deaths during these two months.^4^ The current analysis dissects the validity of this approach using data from different academies and from Nobel laureates.

## METHODS

### Evaluated cohorts

Academician cohorts comprise populations of high socioeconomic status with maximal educational and scholarly achievement – by definition. They are also selected with extreme survival bias, since to be elected in the academies, inducted members typically should survive into advanced age. To estimate the magnitude of selectiveness in terms of age structure and actuarial risk, data were analyzed from CAE (previously used by ref. 4 to estimate excess deaths in Mainland China),^5,6^ the British Academy (BA),^7,8^ the Royal Society,^9,10^ and the Academy of Athens,^11^ national academies that maintain databases of living and deceased members.^5–11^

To obtain comparative information and insights for potential missingness in academies’ data, Nobel laureates^12^ were similarly evaluated. Nobel laureates are another cohort of even more accomplished and privileged individuals. Death information on laurates should be complete given their extremely famous public status, while less strong assurance exists for academies’ data.

### Validity checking for missing death data

To test for potential missing death data in assessed academies, online information was perused on samples of their earliest elected members shown as living in their websites as of March 2024. Online searches with their names aimed to identify their birth year and find any potential obituary or other mention of death.

### Actuarial risk

For cohorts without overt missingness based on the previous step, annual actuarial risks were calculated. For CEA, in analogy to a previous analysis,^4^ the 12-month periods start on December 1, so as to capture in a single 12-month period the impact the COVID-19 wave in December 2022-January 2023; and 5-prepandemic years were also considered (December 2017-November 2022). For BA, only years of birth and death were available and calendar years were considered (2017-2023). For each 12-month period or calendar year, death risk was calculated overall and for the strata of academicians <80, 80-84 and ≥85 year old at the beginning of the time interval. Calendar year death risks (2017-2023) was also calculated for Nobel laureates and for the subset where USA is listed as a country in the Nobel Prize webpage (regardless of other countries are also listed).^13,14^ Annual death risk in the 80-84 and ≥85 strata (that comprise the vast majority of deaths) were compared with general population actuarial risks in China, UK and USA, based on WHO 2019 lifetables for men.^15^ In advanced age strata, these academies and Nobel laureates include very small minorities of women.^5–14^

### Excess death peaks

The ability of the examined cohorts to provide insights about the excess deaths in the general population (or urban-dwelling general population, as assumed in ref. 4) was evaluated by examining the annual deaths in December 2022-November 2023 versus the average of the 5 years that preceded wide SARS-CoV-2 circulation (December 2017-November 2022) in Mainland China; and the average annual deaths during three pandemic years (2020-2022) versus three pre-pandemic years (2017-2019) in the BA and the Nobel laureate cohort (and US subset). Incidence risk ratios were calculated along with 95% confidence intervals (CIs) for the compared periods. For CAE, it is impossible to juxtapose these relative changes against reliable excess death estimates in the general population, since death registration is deficient in Mainland China.^16^ For the UK and US, they were juxtaposed to previously published excess death estimates for 2020-2022 versus the average of 2017-2019 considering weekly death data and changes in population age structure.^17^

### Probe for selective reporting

Previously published^4^ inferences on excess deaths in Mainland China used CAE. A search was made online to examine if other academies in China have available death counts in the periods of interest.

## RESULTS

### Missing death data

Of the first 16 members in alphabetical listing of the inaugural class of 1994 of CAE, the CAE website listed 6/16 as deceased. For another 8, online information provided birth years ranging from 1930 to 1935 and no mention of death was found; for 2 more not even a birth year could be found. In the BA living Fellows list, among the 16 Fellows with the earliest election years, online information listed birth years from 1927 to 1939 without any mention of death. In the Royal Society, among the 19 earliest elected Fellows (elected in 1962-1973) who were listed in the dataset of alive Fellows, 3 were found to have died. For the other 16 (birth years 1923-1939), no death mention was found. Missingness in the Royal Society data was overtly obvious also in the Past Fellows dataset: 32 to 53 annual deaths were recorded between 2016 and 2020, but recorded deaths implausibly declined to 12 in 2021, 5 in 2022, and 0 in 2023 and 2024 (last access, March 18, 2024). In the Academy of Athens, among the 14 earliest elected members (of all types, elected in 1970-1986) who were listed in its website as alive, 5 were found to have died. Therefore, Royal Society and Academy of Athens data were not trusted for in-depth analyses. Naïve calculations of number of deaths in 2020-2022 versus 2017-2019 based on information of their websites would have spuriously suggested 35% relative decrease (70 versus 108 deaths) in the pandemic period for the Royal Society and 32% relative decrease (15 versus 22 deaths) for the Academy of Athens.

### Actuarial risk

In the CEA, annual death rates during the 5 years before December 2022 varied between 1.6% and 2.2% (Table 1). They varied between 1.0% and 2.1% among 80-84 years old CAE members and between 4.4% and 6.4% for those ≥85 years old. According to the WHO life tables for China in 2019, annual actuarial death risk for men in these two age strata in the general population were 11.5% and 26.5%, i.e. 4-12 times higher than CAE members. Even during the 12-month period that witnessed the extensive viral spread, CAE death rates remained 2-to 5-fold lower than those of the general population in 2019 (Table 1).

**Table 1.** Death rates of members of the Chinese Academy of Engineering.

| 12 month period | Members<br>alive at<br>beginning of<br>12-month<br>period | Members<br>80-84 / $\geq 85$<br>years alive<br>at beginning<br>of 12-month<br>period | Deaths in<br>12-month<br>period<br>(%) | Deaths 80-84<br>/ $\geq 85$ years<br>during 12-month period | Death rate<br>among 80-84 / $\geq 85$+<br>years (%) |
| --- | --- | --- | --- | --- | --- |
| 12/2017-11/2018 | 874 | 188 / 180 | 16 (1.8) | 4 / 11 | 2.1 / 6.1 |
| 12/2018-11/2019 | 858 | 199 / 198 | 14 (1.6) | 2 / 10 | 1.0 / 5.1 |
| 12/2019-11/2020 | 919 | 190 / 229 | 19 (2.1) | 3 / 15 | 1.6 / 6.6 |
| 12/2020-11/2021 | 900 | 180 / 252 | 15 (1.7) | 3 / 11 | 1.7 / 4.4 |
| 12/2021-11/2022 | 969 | 166 / 280 | 21 (2.2) | 2 / 18 | 1.2 / 6.4 |
| 12/2022-11/2023 | 948 | 158 / 287 | 45 (4.8) | 4 / 39 | 2.5 / 13.6 |

Data are compiled from references 4,5 and 6 and number of new academicians in each election year are cross-verified from online announcements. New academicians are elected biennially, typically in November or the early days of December and here they are considered as members starting December 1 of the year for consistency (67 new members were elected in 2017, 75 in 2019, 84 in 2021, 74 in 2023).

Annual death rates for BA Fellows were 1.2-2.0% pre-pandemic (i.e. similar to CAE) and 1.7-2.4% during pandemic years. BA is also a cohort of predominantly very elderly people (age data are not publicly available for all Fellows, while age at death in years can be calculated for those deceased). Actuarial death risks for 80-84 and 85+ year old male UK general populations were 6.5% in the 80-84 year old and 15.9% in ≥85 year old people, i.e. about half those of China.

Pre-pandemic annual death rates for Nobel laureates were 3-6% overall and specifically 3.1-5.7% in the US subset (Table 2), i.e. higher that CAE and BA. In the US subset, the number of Nobel laureates and annual deaths in the 80-84 years age stratum were too limited to obtain reliable annual death rates (e.g. in 2023 it was 1/27=3.7% and all years had 0-2 deaths each). Annual death rates in the US subset ≥85 years old strata were 11.4-12.2% pre-pandemic and 5.0-12.8% in pandemic years (average 11.7% versus 8.7%). Actuarial death risks for 80-84 and ≥85 years old male USA general populations in 2019 were 6.6% and 14.7%, i.e. similar to the UK.

**Table 2.**
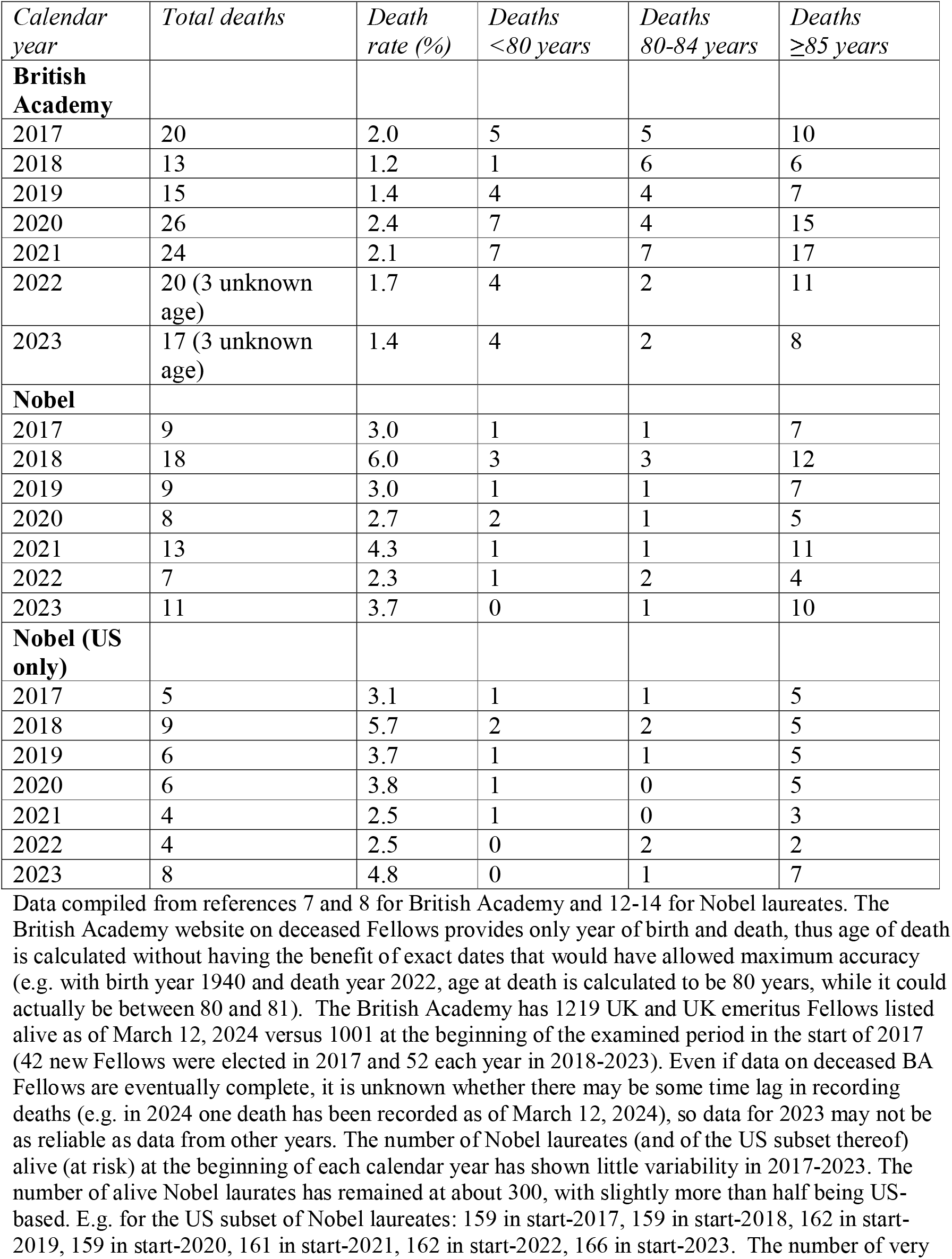
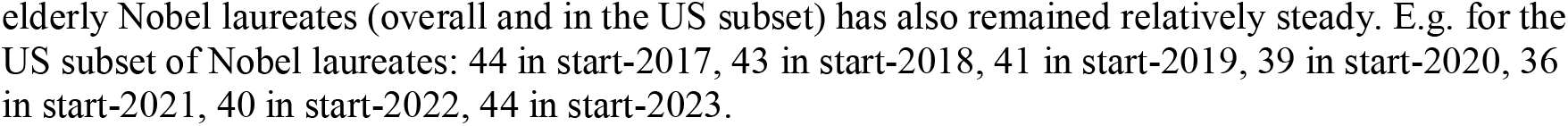
Annual deaths at different age strata recorded among UK Fellows of the British Academy (regular and emeritus), Nobel laureates and the subset of US Nobel laureates.

| *Calendar year* | *Total deaths* | *Death rate (%)* | *Deaths <80 years* | *Deaths 80-84 years* | *Deaths ≥85 years* |
| --- | --- | --- | --- | --- | --- |
| **British Academy** |  |  |  |  |  |
| 2017 | 20 | 2.0 | 5 | 5 | 10 |
| 2018 | 13 | 1.2 | 1 | 6 | 6 |
| 2019 | 15 | 1.4 | 4 | 4 | 7 |
| 2020 | 26 | 2.4 | 7 | 4 | 15 |
| 2021 | 24 | 2.1 | 7 | 7 | 17 |
| 2022 | 20 (3 unknown age) | 1.7 | 4 | 2 | 11 |
| 2023 | 17 (3 unknown age) | 1.4 | 4 | 2 | 8 |
| **Nobel** |  |  |  |  |  |
| 2017 | 9 | 3.0 | 1 | 1 | 7 |
| 2018 | 18 | 6.0 | 3 | 3 | 12 |
| 2019 | 9 | 3.0 | 1 | 1 | 7 |
| 2020 | 8 | 2.7 | 2 | 1 | 5 |
| 2021 | 13 | 4.3 | 1 | 1 | 11 |
| 2022 | 7 | 2.3 | 1 | 2 | 4 |
| 2023 | 11 | 3.7 | 0 | 1 | 10 |
| **Nobel (US only)** |  |  |  |  |  |
| 2017 | 5 | 3.1 | 1 | 1 | 5 |
| 2018 | 9 | 5.7 | 2 | 2 | 5 |
| 2019 | 6 | 3.7 | 1 | 1 | 5 |
| 2020 | 6 | 3.8 | 1 | 0 | 5 |
| 2021 | 4 | 2.5 | 1 | 0 | 3 |
| 2022 | 4 | 2.5 | 0 | 2 | 2 |
| 2023 | 8 | 4.8 | 0 | 1 | 7 |

Data compiled from references 7 and 8 for British Academy and 12-14 for Nobel laureates. The British Academy website on deceased Fellows provides only year of birth and death, thus age of death is calculated without having the benefit of exact dates that would have allowed maximum accuracy (e.g. with birth year 1940 and death year 2022, age at death is calculated to be 80 years, while it could actually be between 80 and 81). The British Academy has 1219 UK and UK emeritus Fellows listed alive as of March 12, 2024 versus 1001 at the beginning of the examined period in the start of 2017 (42 new Fellows were elected in 2017 and 52 each year in 2018-2023). Even if data on deceased BA Fellows are eventually complete, it is unknown whether there may be some time lag in recording deaths (e.g. in 2024 one death has been recorded as of March 12, 2024), so data for 2023 may not be as reliable as data from other years. The number of Nobel laureates (and of the US subset thereof) alive (at risk) at the beginning of each calendar year has shown little variability in 2017-2023. The number of alive Nobel laureates has remained at about 300, with slightly more than half being US-based. E.g. for the US subset of Nobel laureates: 159 in start-2017, 159 in start-2018, 162 in start-2019, 159 in start-2020, 161 in start-2021, 162 in start-2022, 166 in start-2023. The number of very
elderly Nobel laureates (overall and in the US subset) has also remained relatively steady. E.g. for the US subset of Nobel laureates: 44 in start-2017, 43 in start-2018, 41 in start-2019, 39 in start-2020, 36 in start-2021, 40 in start-2022, 44 in start-2023.

### Excess death peaks

In CEA (Table 3), the relative increase in death rate during the 12-month period after wide SARS-CoV-2 circulation started was very high (+158%, 95% CI 80-269%). Most pandemic deaths happened in the ≥85 years age stratum (relative death rate pandemic increase +158%, 95% CI 78-274%). The CEA cohort became older over time (Table 1).

**Table 3.**
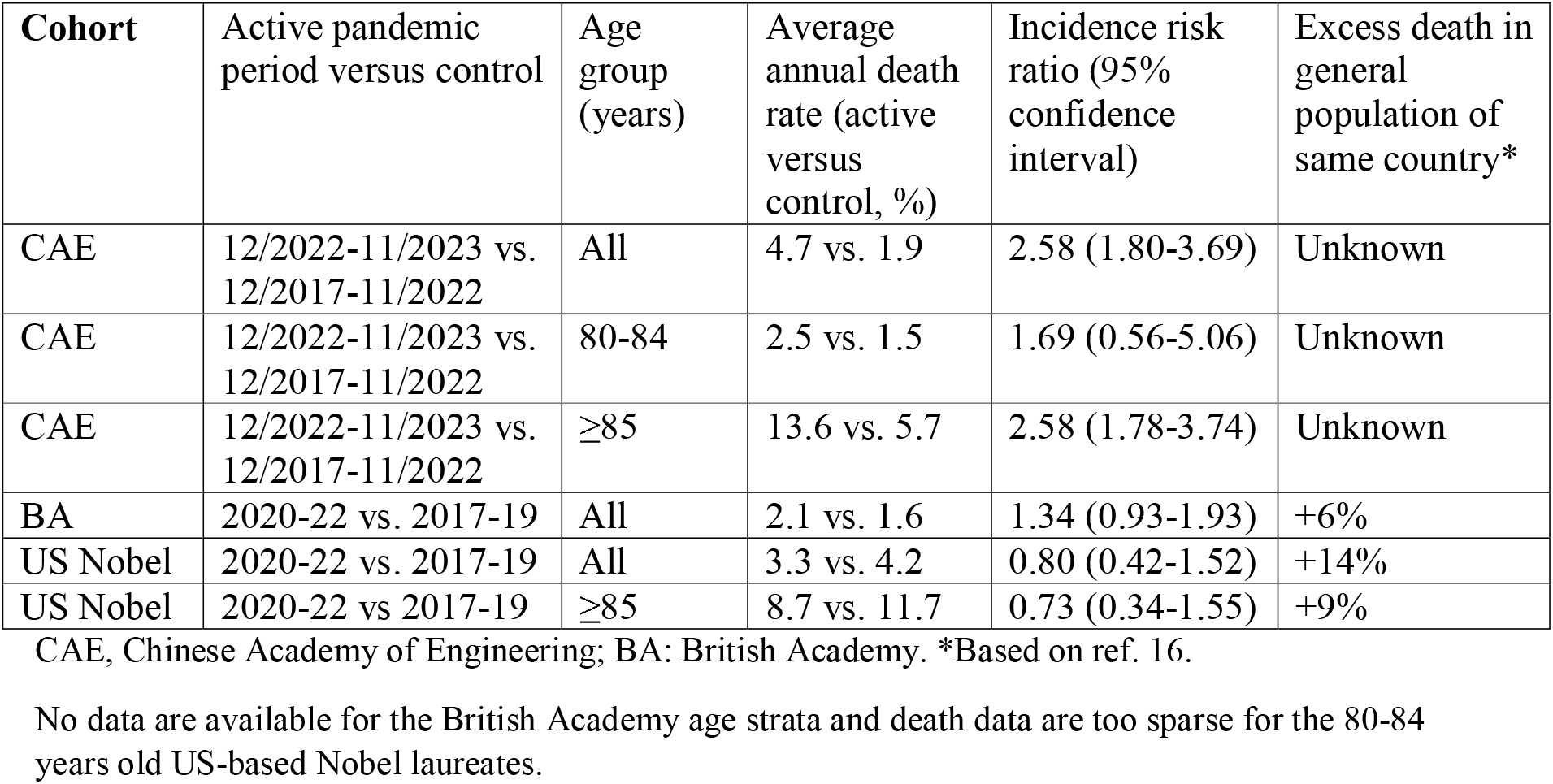
Death rates in active pandemic period versus control period, calculated incidence risk ratio and juxtaposed excess death estimates.

In BA (Table 3), the relative increase in the death rate in 2020-2022 versus 2017-2019 was 34% (95% CI -7% decrease to 93% increase). Data are unavailable to generate age strata estimates. UK general population excess deaths in 2020-2022 were 6% overall, and smaller in the elderly (e.g. 4% for ≥85 years old).

Nobel laureates (Table 3) had no discernible death peak during the pandemic. A modest peak actually occurred in 2018. Annual deaths tended to be lower during the pandemic years than pre-pandemic. The estimated relative decrease was -20% (95% CI, -58% decrease to 52% increase) in the US subset overall and -27% among its ≥85 years old laureates (95% CI, -66% decrease to 55% increase). Conversely, USA national data showed 14% excess deaths overall and 9% among those ≥85 years old.

### Selective reporting

Mainland China has three major national academies (CAE, Chinese Academy of Sciences (CAS), Chinese Academy of Social Sciences (CASS)). No information on birth and death years was listed by CASS. Living members data were available for CAS,^18^ but the CAS deceased members webpage was defunct as of March 12, 2024. Nevertheless, both CAE and CAS death data were presented in the New York Times story.^3^ CAE and CAS have approximately the same number of members (currently 968 versus 866). CAS has slightly lower average age, larger proportion of members ≥95 years old (probably because of its earlier launch year) and substantially lower portion of members 90-94 years old than CAE. Therefore, expected number of deaths would be slightly higher in CAE than in CAS. However, there were far more deaths of CAE members than CAS members in the window of maximal interest (December 2022 to January 2023): 28 versus 12 (or 26 versus 11 when limited to deaths ≥80 years old, as focused in ref. 4).^3^

## DISCUSSION

Cohorts of academicians are, by definition, subject to extreme selection bias and cover a tiny proportion of the general population age structure. Extrapolating to the whole general population, in particular younger strata, can thus be highly misleading. Empirical data from BA and Nobel laureates also show that trends for both increases (BA) and decreases (Nobel laureates and their US subset) were seen during the pandemic in the death rates of these prestigious people. These point estimates were far from the annual excess deaths in the general population of the respective country. BA Fellows’ data point estimate would suggest roughly 5-fold (or more) higher excess deaths than what really happened in the general population, while inspecting Nobel laureate data naively would give the totally false impression that a peak, if any, might had occurred in 2018, i.e. before the pandemic. Given that all these cohorts have limited membership, the sample size is small and thus 95% CIs are very wide. Given the potential biases involved, uncertainty is even larger than what the already impressive 95% CIs would suggest. The bias magnitude may vary across countries, settings, and academician cohorts. Thus, using such data to infer excess deaths in countries where excess deaths cannot be otherwise calculated in the population directly from reliable death registration, could lead to severe miscalculations.

Several hundreds of honorific academies exist. The major focus in public media on the Chinese academies may herald selective reporting bias. Differences seemed too stark even to non-scientist journalists^3^ and thus worth writing a story about them. Modest or no differences in death rates (or even lower death rates during the pandemic versus pre-pandemic years, as among Nobel laureates) would not have attracted journalistic interest. Interestingly, selective analysis reporting bias was apparently worsened in the scientific peer-reviewed literature versus the original journalistic investigation media coverage. E.g. the analysis of ref. 4 included only CAE data, while the New York Times article^3^ that inspired that analysis (as the authors admit)^4^ had presented data on both CAE and CAS. Excess death estimates using CAS might have been only about half those calculated using CAE data.

The combined effect of diverse biases cannot be generalized across all academy cohorts. In the specific example where excess deaths in Mainland China were estimated,^4^ the most likely scenario is large over-estimation in the published calculations. Comparing cohorts of academicians to the urban-dwelling subset of the general population (as in ref. 4), diminishes this bias modestly. E.g. in Mainland China, there is a rural-urban life expectancy gap; however, this gap is comparatively modest (1-2 years),^19^ thus most of the bias remains.

Excess death inferences using academician cohorts can be further markedly distorted, if the academies’ datasets do not capture all deaths and if birth/death dates have errors. While the latter error is hopefully uncommon for cohorts of famous people, there is no guarantee that academies can keep perfect track of all their members’ deaths. The very low death rates described in the Chinese and British academies analyzed here may be partly explained also by unreported deaths. No overt cases of missed deaths were documented for CAE and BA, at least in the fashion that missingness could be overtly documented for Royal Society and Academy of Athens datasets. However, it is tantalizing that death rates in CAE and BA were substantially lower than those of Nobel laureates who are likely to be at least as protected and privileged healthcare-wise. This may also reflect missing deaths in CAE and BA.

Extrapolating from very elderly cohorts poses further challenges. E.g., excess death calculations for all Mainland China based on CAE^4^ used death data on those ≥80 years old and extrapolated to younger populations using also data from Hong Kong where death registration is apparently more complete. In Mainland China, people ≥80 years old represent only 2.5% of the population.^20^ In fact, recorded deaths in academicians aged 80-84 years were too few; unsurprisingly the wide CI of the excess deaths’ estimate in that age stratum included even a possibly large death deficit rather than excess.^4^ Academician data offer mostly information on deaths in the stratum of ≥85 years old, but only ∼1% of the general population in China is ≥85 years old.^20^ Different countries where death registration data are reliably complete have shown variable rates of excess deaths across age strata.^17^ On average, excess death (expressed in percentages above expected deaths, p%) have been higher in the elderly than in non-elderly strata; particularly in France, Slovenia, and Poland, these differences have been stark.^17^ Conversely, a few countries (USA, Canada, UK and Chile) have shown higher p% in the nonelderly, probably boosted by non-COVID-19 causes during the pandemic (e.g. increased overdose deaths or deaths from poor access to care).^1,21,22^ Also during the pandemic there were no excess deaths but consistently reduced mortality for children and adolescents.^17,23^ Inappropriate extrapolation of the mortality experience from elderly frail populations to non-frail and younger populations was a major misunderstanding perpetuated in various forms throughout the pandemic.^24–27^

Given the selective privileged status, cohorts of academicians probably include several people with extreme frailty, who are nevertheless maintained alive because of access to excellent care. Peak death burden during pandemic waves may be modulated from mortality displacement (harvesting) effects, if previous years have seen lower or higher than average death rates. Mortality displacement has been documented at a seasonal timeframe (e.g. inverse coupling of summer heat-related deaths and winter deaths among frail individuals)^28,29^ or longer timeframes, as described previously specifically also with COVID-19.^30^ However, in contrast to other frail populations (e.g. cohorts of long-term care residents) where mortality displacement could be readily documented within months,^31^ academician cohorts are very heterogeneous. Heterogeneity cannot be characterized without concomitant health data on the members. Moreover, lacking such data, one cannot adjust for comorbidities and frailty.

Besides data from academicians, other cohorts related to universities have been used for excess death calculations. Xiao et al.^32^ used obituaries from 3 Chinese universities. Populations of university professors and staff are also select groups with higher than average socioeconomic status. Thus they may still be privileged in life expectancy versus the general population, but the advantage is probably less pronounced than for prestigious academy members. Therefore, similar considerations, albeit to a smaller extent, apply to such cohorts. Moreover, missingness may be more prominent in university cohorts, since it is very unlikely that all deaths are captured through obituary collections. Worse, missingness may be uneven over time, with heightened interest in obituaries (and thus more obituaries written and captured) during a crisis or perceived crisis – as during active pandemic waves. Using obituaries from a wider population^32,33^ makes the sample less selective, but missingness (and uneven missingness over time) may become even more prominent. Reliability may be higher in sharply defined populations with focused, exclusive venues for publishing obituaries, e.g. Amish and Mennonites.^34,35^ Using information from online searches for funeral services and related terms^32,33^ expands further the data collection, but the representativeness of collected samples is difficult to gauge. Moreover, online searches are probably affected by both real and perceived crises and it is difficult to separate the two. E.g. if there is a perception (reinforced by media, social media, and or public health announcements) of a major lethal problem, more people may search online with terms related to that problem. Thus these methods would tend to generate inflated estimates of excess deaths in the general population. These estimates are accordingly higher than those obtained by standardizing population structure and infection fatality rates against those of populations with accurate death registration.^36,37^

Overall, proper death registration is indispensable for estimating excess deaths. In its absence, well-characterized cohorts may be meaningfully used, provided their participants are representative of the general population - or can be properly adjusted to be representative thereof - and information on deaths is complete. One should avoid using cohorts that may suffer from major biases.

## Data Availability

All key data are in the manuscript and in databases that are already open to the public.

## REFERENCES

1. Ioannidis JPA, Zonta F, Levitt M. Flaws and uncertainties in pandemic global excess death calculations. Eur J Clin Invest. 2023 Aug;53(8):e14008. doi: 10.1111/eci.14008.

2. Nepomuceno MR, Klimkin I, Jdanov DA, Alustiza-Galarza A, Shkolnikov VM. Sensitivity Analysis of Excess Mortality due to the COVID-19 Pandemic. Popul Dev Rev. 2022 Jun;48(2):279–302. doi: 10.1111/padr.12475.

3. New York Times (February 3, 2023), https://www.nytimes.com/interactive/2023/02/05/world/asia/china-obits-covid.html, last accessed March 11, 2024.

4. Raphson L, Lipsitch M. Estimated excess deaths due to COVID-19 among the urban population of Mainland China, December 2022 to January 2023. Epidemiology. 2024 Jan 29. doi: 10.1097/EDE.0000000000001723.

5. Chinese Academy of Engineering, list of members, https://en.cae.cn/cae/html/en/col2229/column_2229_1.html, last accessed March 10, 2024.

6. Chinese Academy of Engineering, list of deceased members, https://www.cae.cn/cae/html/main/col56/column_56_1.html, last accessed March 10, 2024.

7. British Academy. Fellows of the British Academy, https://www.thebritishacademy.ac.uk/fellows/?order=year_elected#filtered-list, last accessed March 10, 2024.

8. British Academy. List of deceased Fellows, https://www.thebritishacademy.ac.uk/deceased-fellows/, last accessed March 10, 2024.

9. Royal Society. List of Fellows, https://royalsociety.org/fellows-directory/, last accessed March 12, 2024.

10. Royal Society. Search Past Fellows, https://catalogues.royalsociety.org/CalmView/personsearch.aspx?src=CalmView.Persons, last accessed March 18, 2024.

11. List of members of Academy of Athens, http://www.academyofathens.gr/el/members, last accessed March 18, 2024.

12. Database of Nobel Prize laureates, https://public.opendatasoft.com/explore/dataset/nobel-prize-laureates/table/?flg=en-us&disjunctive.category, last accessed March 14, 2024.

13. All Nobel Prizes, https://www.nobelprize.org/prizes/lists/all-nobel-prizes/, last accessed March 14, 2024.

14. List of Nobel laureates by country, https://en.wikipedia.org/wiki/List_of_Nobel_laureates_by_country, last accessed March 14, 2024.

15. WHO. Global health observatory data repository. Life tables by country. https://www.who.int/data/gho/data/indicators/indicator-details/GHO/gho-ghe-life-tables-by-country, last accessed March 10, 2024

16. Zeng X, Adair T, Wang L, Yin P, Qi J, Liu Y, Liu J, Lopez AD, Zhou M. Measuring the completeness of death registration in 2844 Chinese counties in 2018. BMC Med. 2020 Jul 3;18(1):176.

17. Ioannidis JPA, Zonta F, Levitt M. Variability in excess deaths across countries with different vulnerability during 2020-2023. Proc Natl Acad Sci U S A. 2023 Dec 5;120(49):e2309557120. doi: 10.1073/pnas.2309557120.

18. Members of Chinese Academy of Sciences, http://english.casad.cas.cn/mem/divsions/domap/, last accessed March 10, 2024.

19. Han S, Su B, Zhao Y, Chen C, Zheng X. Widening rural-urban gap in life expectancy in China since COVID-19. BMJ Glob Health. 2023 Sep;8(9):e012646. doi: 10.1136/bmjgh-2023-012646.

20. China population pyramid 2023, https://www.populationpyramid.net/china/2023/, last accessed March 8, 2024.

21. https://www.cdc.gov/nchs/pressroom/nchs_press_releases/2022/202205.htm, last accessed March 12, 2024.

22. Nadarajah R, Wu J, Hurdus B, Asma S, Bhatt DL, Biondi-Zoccai G, Mehta LS, Ram CVS, Ribeiro ALP, Van Spall HGC, Deanfield JE, Lüscher TF, Mamas M, Gale CP. The collateral damage of COVID-19 to cardiovascular services: A meta-analysis. Eur Heart J. 2022;43:3164–3178.

23. GBD 2021 Demographics Collaborators. Global age-sex-specific mortality, life expectancy, and population estimates in 204 countries and territories and 811 subnational locations, 1950–2021, and the impact of the COVID-19 pandemic: a comprehensive demographic analysis for the Global Burden of Disease Study 2021. Lancet 2024; March 11, 2024, 10.1016/S0140-6736(24)00476-8.

24. Axfors C, Ioannidis JPA. Infection fatality rate of COVID-19 in community-dwelling elderly populations. Eur J Epidemiol. 2022 Mar;37(3):235–249. doi:10.1007/s10654-022-00853-w.

25. Pezzullo AM, Axfors C, Contopoulos-Ioannidis DG, Apostolatos A, Ioannidis JPA. Age-stratified infection fatality rate of COVID-19 in the non-elderly population. Environ Res. 2023 Jan 1;216(Pt 3):114655. doi:10.1016/j.envres.2022.114655. Epub 2022 Oct 28.

26. Eigl ES, Widauer SS, Schabus M. Burdens and psychosocial consequences of the COVID-19 pandemic for Austrian children and adolescents. Front Psychol. 2022 Nov 11;13:971241. doi: 10.3389/fpsyg.2022.971241.

27. Schabus, M., Eigl, E.-S., and Widauer, S. S. (2022). The coronavirus pandemic: Psychosocial burden, risk-perception, and attitudes in the Austrian population and its relation to media consumption. Front. Public Health 10:921196. doi: 10.3389/fpubh.2022.921196.

28. Rocklöv J, Forsberg B, Meister K. Winter mortality modifies the heat-mortality association the following summer. Eur Respir J. 2009;33(2):245–51.

29. Lytras T, Pantavou K, Mouratidou E, Tsiodras S. Mortality attributable to seasonal influenza in Greece, 2013 to 2017: variation by type/subtype and age, and a possible harvesting effect. Euro Surveill. 2019 Apr;24(14):1800118.

30. Zahl PH, Hemström Ö, Johansen R, Mamelund SE. Mortality in Norway and Sweden during the COVID-19 pandemic 2020-22: A comparative study. J Infect Public Health. 2023 Nov 3:S1876–0341(23)00371-4.

31. Ballin M, Ioannidis JP, Bergman J, Kivipelto M, Nordström A, Nordström P. Time-varying risk of death after SARS-CoV-2 infection in Swedish long-term care facility residents: a matched cohort study. BMJ Open. 2022 Nov 24;12(11):e066258.

32. Xiao H, Wang Z, Liu F, Unger JM. Excess All-Cause Mortality in China After Ending the Zero COVID Policy. JAMA Netw Open. 2023 Aug 1;6(8):e2330877. doi: 10.1001/jamanetworkopen.2023.30877.

33. Huang L, Li OZ, Yin X. Inferring China’s excess mortality during the COVID-19 pandemic using online mourning and funeral search volume. Sci Rep. 2023 Sep 20;13(1):15665. doi: 10.1038/s41598-023-42979-1.

34. Eash-Scott D, Stoltzfus D, Brenneman R. "The Graves Cannot Be Dug Fast Enough": Excess Deaths Among US Amish and Mennonites During the 1918 Flu Pandemic. J Relig Health. 2024 Feb;63(1):652–665. doi: 10.1007/s10943-023-01899-0.

35. Stein RE, Corcoran KE, Colyer CJ, Mackay AM, Guthrie SK. Closed but Not Protected: Excess Deaths Among the Amish and Mennonites During the COVID-19 Pandemic. J Relig Health. 2021 Oct;60(5):3230–3244. doi: 10.1007/s10943-021-01307-5.

36. Ioannidis JPA, Zonta F, Levitt M. Estimates of COVID-19 deaths in Mainland China after abandoning zero COVID policy. Eur J Clin Invest. 2023 Apr;53(4):e13956. doi: 10.1111/eci.13956.

37. Ioannidis JPA, Zonta F, Levitt M. What Really Happened During the Massive SARS-CoV-2 Omicron Wave in China? JAMA Intern Med. 2023 Jul 1;183(7):633–634. doi: 10.1001/jamainternmed.2023.1547.

